# Comparison of Mechanical Tissue Properties Using MyotonPRO and Time-Harmonic Elastography: Understanding Fundamental Differences and Statistical Relationships

**DOI:** 10.64898/2026.05.20.26353658

**Authors:** Eduard Kurz, Giacomo Valli, Tom Meyer, Stefan Pröger, René Schwesig, Thomas Bartels, Karl-Stefan Delank, Ingolf Sack, Hossein S. Aghamiry

## Abstract

**Purpose:** MyotonPRO (MTP) and time-harmonic elastography (THE) are increasingly used to assess muscle mechanical properties, yet they operate on fundamentally different physical principles. MTP measures composite MTP stiffness (N/m) through surface oscillations, while THE quantifies intrinsic shear modulus (THE stiffness, kPa) via propagating shear waves. This study aimed at systematically compare MTP and THE measurements in the vastus lateralis muscle across different contraction intensities and examine how the skin layer and subcutaneous fat (SLSF) thickness influence their relationship.

**Methods:** Twenty-six healthy adults (15 males, 11 females; age 25 [SD 4] years) underwent MTP and THE measurements of the vastus lateralis at rest and during isometric contractions at 15% and 30% maximal voluntary contraction (MVC). Effects of contraction intensities on tissue properties were assessed using univariate analyses of variance with repeated measures. Associations between the different outcomes of THE and MTP technologies were explored using Pearson’s correlations and partial correlation coefficients separately for each contraction intensity with adjustment of the SLSF thickness of participants.

**Results:** Both technologies detected contraction intensity-dependent stiffening across all outcomes (p < 0.001). THE stiffness increased from 5.3 [1.2] kPa at rest to 15.6 [6.1] kPa at 30% MVC; THE wave attenuation increased from 0.83 [0.19] to 1.42 [0.36] s/m while MTP stiffness increased from 337.3 [49.3] N/m at rest to 529.4 [160.7] N/m at 30% MVC. Correlations between modalities were weak and condition-dependent. THE wave attenuation did not significantly correlate with any MTP outcome across conditions.

**Conclusion:** MTP and THE detect contraction-induced stiffening through fundamentally different physical mechanisms and should not be regarded as interchangeable. Their correlation is modest at rest and breaks down (or reverses) during active contraction, with subcutaneous fat as a key modifying factor.

Clinical trial number: Not applicable.

## Introduction

Measuring muscle stiffness with non-invasive methods has become increasingly important. It is useful for understanding neuromuscular function, tracking training adaptations, and diagnosing pathologies. Two technologies have emerged as popular tools: surface mechanical devices like MyotonPRO (MTP) and elastography techniques like time-harmonic elastography (THE). While both claim to measure tissue “stiffness”, they operate on fundamentally different physical principles, and their outputs cannot be directly converted without substantial approximations and assumptions.

MTP has become widely used in clinical and sports settings because it is portable, easy to use, and gives quick results [1, 2]. It works by tapping the tissue and analyzing how it oscillates. The processing pipeline essentially treats the probe-tissue system as a damped spring (see Appendix A1). THE takes a different approach: it uses continuous vibration to send shear waves through the tissue, and the speed of these waves reveals the intrinsic stiffness of tissues [3, 4] (see Appendix A2).

Despite how widely both methods are used, only a few studies have directly compared them. Feng et al. [5] examined the relationship between shear wave elastography and MTP in muscle and tendon, finding moderate correlations (r = 0.40−0.60) but substantial discrepancies between the actual values. Their work suggested that while both methods detect changes in tissue mechanics, they are measuring different things and cannot be directly converted without accounting for fundamental differences in what they actually measure.

A central challenge in comparing these technologies is that they measure fundamentally different physical quantities. MTP yields MTP Stiffness in N/m, which depends on both the material properties and the geometry of what is measured (Figure 1). Elastography yields shear modulus in kPa, an intrinsic material property that does not depend on geometry [6, 7]. They also work at different frequencies: MTP characterizes tissue at its natural resonance (10–50 Hz) while THE uses controlled frequencies (60–80 Hz). Soft biological tissue such as skeletal muscle behaves differently at different frequencies [8, 9]. Furthermore, MTP integrates the mechanical response from the surface to approximately 15–20 mm depth (skin, fat, fascia, muscle) while THE can isolate specific tissue layers [4, 10].

**Figure 1.**
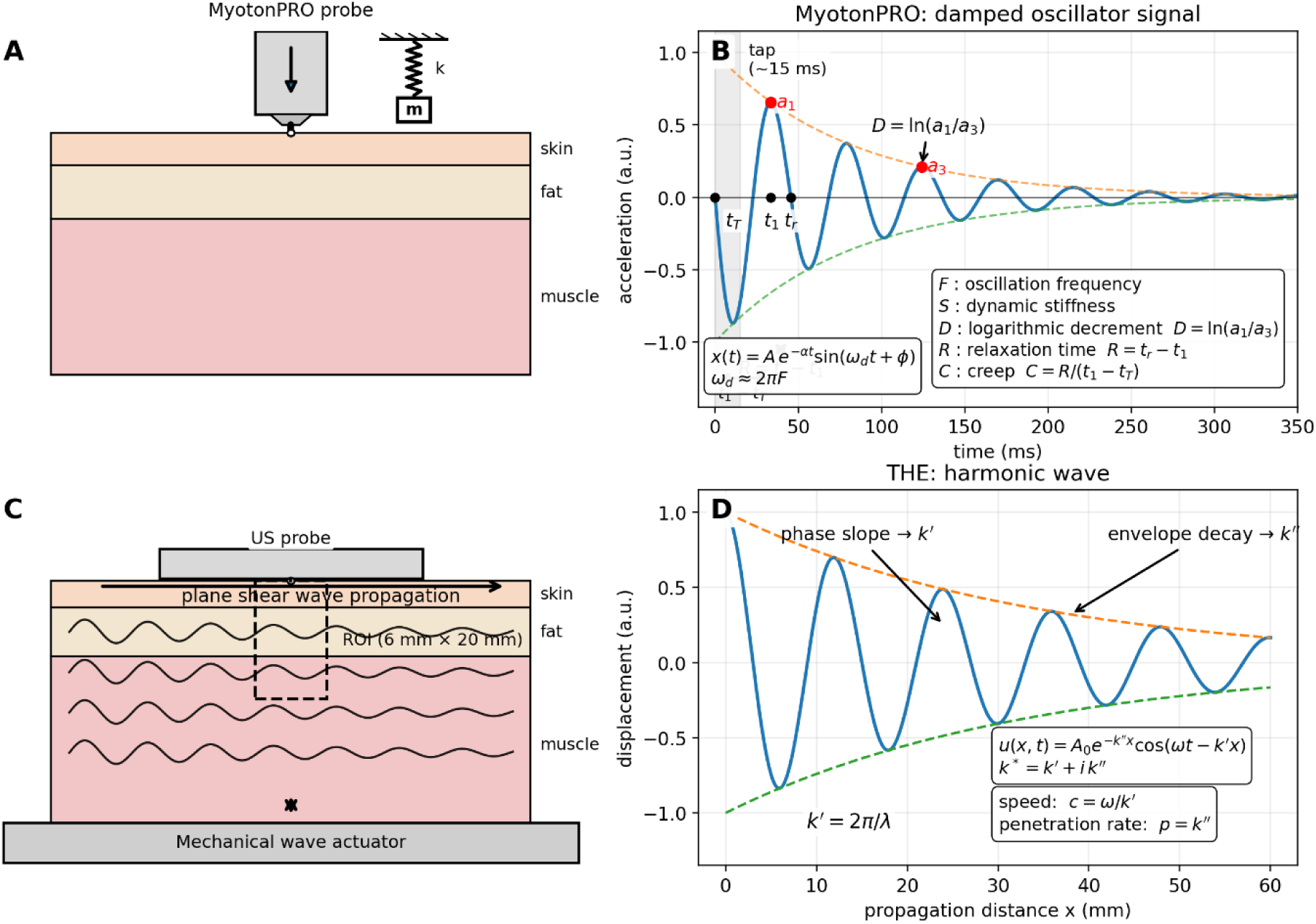
(A) MyotonPRO measurement concept: a brief mechanical tap is applied at the skin surface, and the tissue response is modeled as a damped mass-spring system (spring constant *k*, effective mass *m*), with the probe tip in contact with the skin. (B) Example MyotonPRO time response illustrating parameter extraction: successive positive peaks *a*_1_and *a*_3_define the logarithmic decrement *D* = ln (*a*_1_/*a*_3_); relaxation time is estimated from *R* = *t*_*r*_ − *t*_1_, where *t*_1_ is the time of the first peak and *t*_*r*_ is the subsequent zero-crossing; the creep index is computed as *C* = R/(*t*_1_ − *t*_*T*_), with *t*_*T*_ denoting the tap onset. (C) Time-harmonic elastography (THE) setup schematic: a mechanical actuator excites shear waves in layered tissue (skin-fat-muscle), while an ultrasound probe tracks wave motion. (D) Harmonic shear-wave model used in THE, *u*(x, t) = *A*_0_*e*^−*k*′′*x*^cos (𝜔*t* − *k*^′^*x*), where the complex wavenumber *k*^∗^ = *k*^′^ + i*k*^′′^ provides wave speed *c* = ω/*k*^′^ and attenuation 𝛼 = ω/*k*^′′^. The THE ROI used in this study for comparison with MyotonPRO is shown in panel C.

This has direct clinical relevance, as these technologies are frequently used interchangeably or assumed to measure the same quantity, which can lead to misinterpretation of results. Furthermore, determining whether the two methods provide complementary or redundant information would inform the selection of the appropriate tool for a given application.

Based on the background presented, the goal is to systematically compare outcomes of MTP and THE in the vastus lateralis muscle at rest (0% MVC) and during low-level isometric contractions (15% and 30% MVC). We expected both methods to detect contraction intensity-dependent stiffening, but not in a directly proportional way due to their fundamental differences. We also examined whether anatomical factors such as skin layer and subcutaneous fat (SLSF) thickness modify the agreement between outcomes of the two technologies. The goal is to provide practical guidance for interpreting and comparing results from these widely used muscle assessment tools.

## Materials & Methods

### Study Design & Ethics

This quasi-experimental cross-sectional study was conducted according to the guidelines of the Declaration of Helsinki. The study protocol was approved by the Institutional Review Board at the Medical Faculty of the Martin-Luther-University Halle-Wittenberg (approval number: 2024-208). Written informed consent was obtained from all participants prior to enrollment.

### Participants

Adults of both sexes who were of consent age, had no musculoskeletal injuries, and had no neuromuscular disorders were included. Exclusion criteria were an acute medical history, injuries of the lower extremity, restricted mobility of the knee joints, and any secondary issues that limited the ability of participant to execute the required task. Participants were undergraduate medical or sport science students recruited on a voluntary basis at the local University.

### Experimental Setup & Data Acquisition

Knee extension force (SM-2000N, Interface, Inc., Scottsdale, Arizona, USA) was measured isometrically at the right lower limb. Measurements were made over the distal portion of the vastus lateralis muscle at rest and during force-matching trapezoidal ramp contractions with 12-second plateaus at 15% and 30% of the maximal voluntary contraction (MVC) force (Figure 2). After a brief warm-up at 15% and 30% of the estimated maximal force (M: 6, F: 4 Newtons per kg body mass) including familiarization with the ramp contractions two MVC trials were completed. The maximal force achieved served as reference for the submaximal contraction levels. The sequence was predefined and started with the lowest intensity (rest, 0% MVC) performed before the warm-up protocol followed by the MVC trials and the submaximal contraction levels.

**Figure 2.**
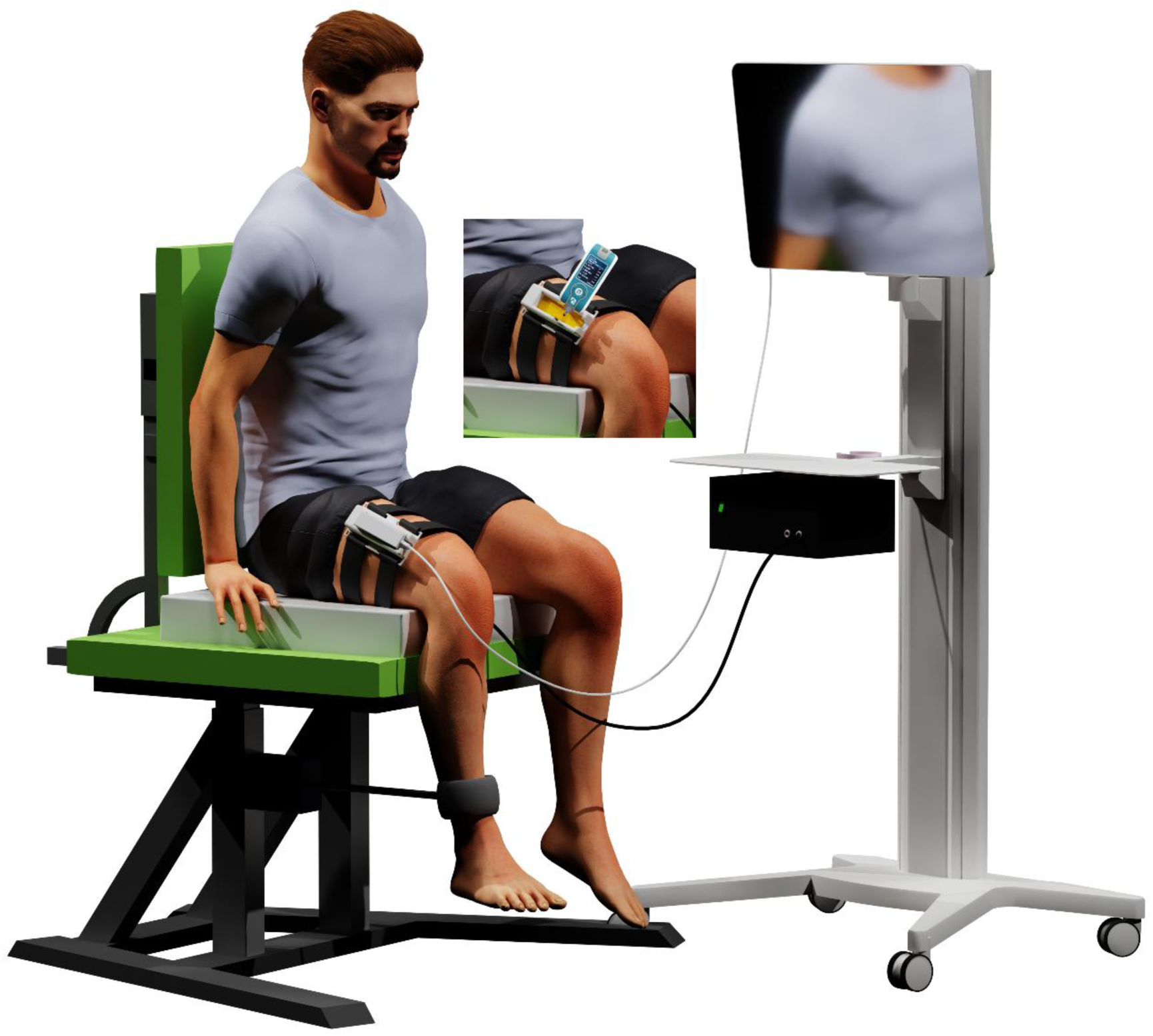
Measurement setup with the participant sitting on a custom-made pillow with integrated loudspeakers and a specifically designed ultrasound probe holder over the vastus lateralis muscle of the right side.

### Data Processing & Outcomes

The THE processing pipeline followed established protocols [4, 11]. Radiofrequency (RF) ultrasound data, where RF denotes the beamformed acquired data, were recorded at 100 frames/s (*F*_*s*_ = 100 *Hz*) for 12 seconds per measurement and underwent four main processing steps: (1) monogenic phase-based displacement estimation using a multiscale optical-flow solver with spatial Tikhonov regularization and truncated-SVD denoising (ranks 2–100) along time coordinate to extract vertical tissue motion between consecutive frames [12, 13]; (2) harmonic isolation using third-order Butterworth filters (60/70/80 Hz ± 1.5 Hz) with controlled-aliasing reconstruction to recover frequency components from the 100 Hz frame rate; (3) directional filtering in the 2-D wavenumber domain using angular Gaussian windows (N=4 directions, σ=π/2, FWHM≈212°) centered at 0° and 180° to emphasize horizontally propagating shear waves while suppressing boundary reflections; and (4) k-MDEV inversion computing local phase gradients to generate shear wave speed (SWS) maps with 1 mm spatial resolution as *c* = ω/*k*^′^ expressed in m/s (Figure 1). The shear modulus G (in kPa or Pa) is computed from the measured SWS via the relationship G = ρ·SWS², where ρ represents tissue mass density. For soft biological tissues, ρ is conventionally assumed to be 1000 kg/m³. The resulting shear modulus provides a material property that reflects the resistance of tissue to shear deformation, independent of geometric factors such as tissue thickness or probe contact area.

Another output of THE is the penetration rate, which quantifies the loss of shear wave energy during propagation through tissue. It is derived from the imaginary component of the complex wavenumber extracted during k-MDEV inversion as *p* = ω/*k*^′′^, and expressed in m/s (Figure 1). The inverse of the penetration rate is a proxy of attenuation. To facilitate comparison with MTP viscosity outcomes, we report the inverse of the THE penetration rate. Higher attenuation indicates greater energy dissipation and is associated with increased tissue viscosity, structural heterogeneity, and interstitial fluid redistribution.

MTP measurements were applied with the probe set perpendicular to the skin surface in multiscan mode (five repetitions). The probe had a preload of 0.18 N, a tap time of 15 ms, and a tap interval of 0.8 s. All repetitions were used to generate one average value per outcome at each contraction intensity. MTP quantifies tissue mechanical properties by analyzing the damped oscillatory response following a brief mechanical impulse. The underlying physical principle treats the probe-tissue system as a damped mass-spring oscillator, where the tissue provides both elastic restoring forces (spring stiffness *k*) and viscous energy dissipation (damping coefficient *c*). Following impulse delivery, the effective mass (*m* ≈ 18 g, comprising the probe and coupled tissue mass) oscillates at the natural frequency of tissue, with amplitude decaying exponentially due to internal friction and viscous damping.

The measured acceleration signal reflects the natural mechanical response of the tissue at its resonant frequency (typically 10–50 Hz for soft tissues), which depends on both tissue stiffness and the oscillating mass. The primary MTP outputs (oscillation frequency *F* and stiffness *S*) are directly extracted from this free vibration response [1, 2, 14]. Importantly, MTP measures the *MTP stiffness* of the probe-tissue system (units: N/m), which represents a composite property integrating contributions from all tissue layers within the measurement depth (approximately 15–20 mm), including skin, subcutaneous fat, fascia, and muscle [15, 16]. All outcomes provided by the MTP device (MTP stiffness, oscillation frequency, logarithmic decrement, relaxation time, creep), as shown in Figure 1, were used for comparisons with THE stiffness as well as THE wave attenuation.

In general, THE provides spatially resolved maps of biomechanical parameters (such as SWS and attenuation) from which values within a specific region of interest (ROI) can be selected. Because the goal here is to compare MTP with THE while integrating contributions from all tissue layers within the measurement depth, we defined the ROI in THE as extending from the tissue surface to a depth of 20 mm, with a width of 6 mm centered beneath the ultrasound probe (i.e., the location where the MTP measurements were performed).

### Data Analysis & Statistics

Data are presented as mean with standard deviation (SD) values. Prior to any inference statistical analysis, the data and residuals distribution were checked visually and using the Shapiro-Wilk test. Normal distribution of the data was confirmed by both methods. Sex differences were explored using Student’s t test for independent samples with Cohen’s d values. Effects of contraction intensities on tissue properties were assessed using univariate analyses of variance with repeated measures (contraction intensity) separately for each outcome and technology. Effect size (ES) values are reported as partial eta-squared. Associations between the different outcomes of THE (2) and MTP (5) technologies were explored using Pearson’s product moment correlations and partial correlation coefficients with adjustment of participants’ SLSF thickness separately for each contraction intensity.

## Results

### Participant Characteristics

Twenty-six healthy adults (M/F: 15/11, age: 25 [SD 4] years, body mass index: 23.8 [2.7] kg/m²) were examined. Each enrolled participant successfully completed all measurements. No participant withdrew their consent. Female participants showed lower relative knee extension torque values than males (M: 5.01 [0.86] Nm/kg, F: 4.27 [0.47] Nm/kg, p = 0.016, Cohen’s d = –1.0). In contrast, male participants revealed a lower SLSF thickness over the vastus lateralis muscle compared with females (M: 5.5 [2.6] mm, F: 8.9 [3.2] mm, p = 0.006, Cohen’s d = 1.2). The strength of the linear relationships between SLSF thickness and tissue stiffness estimations of both technologies at different contraction intensities are displayed in Figure 3.

**Figure 3.**
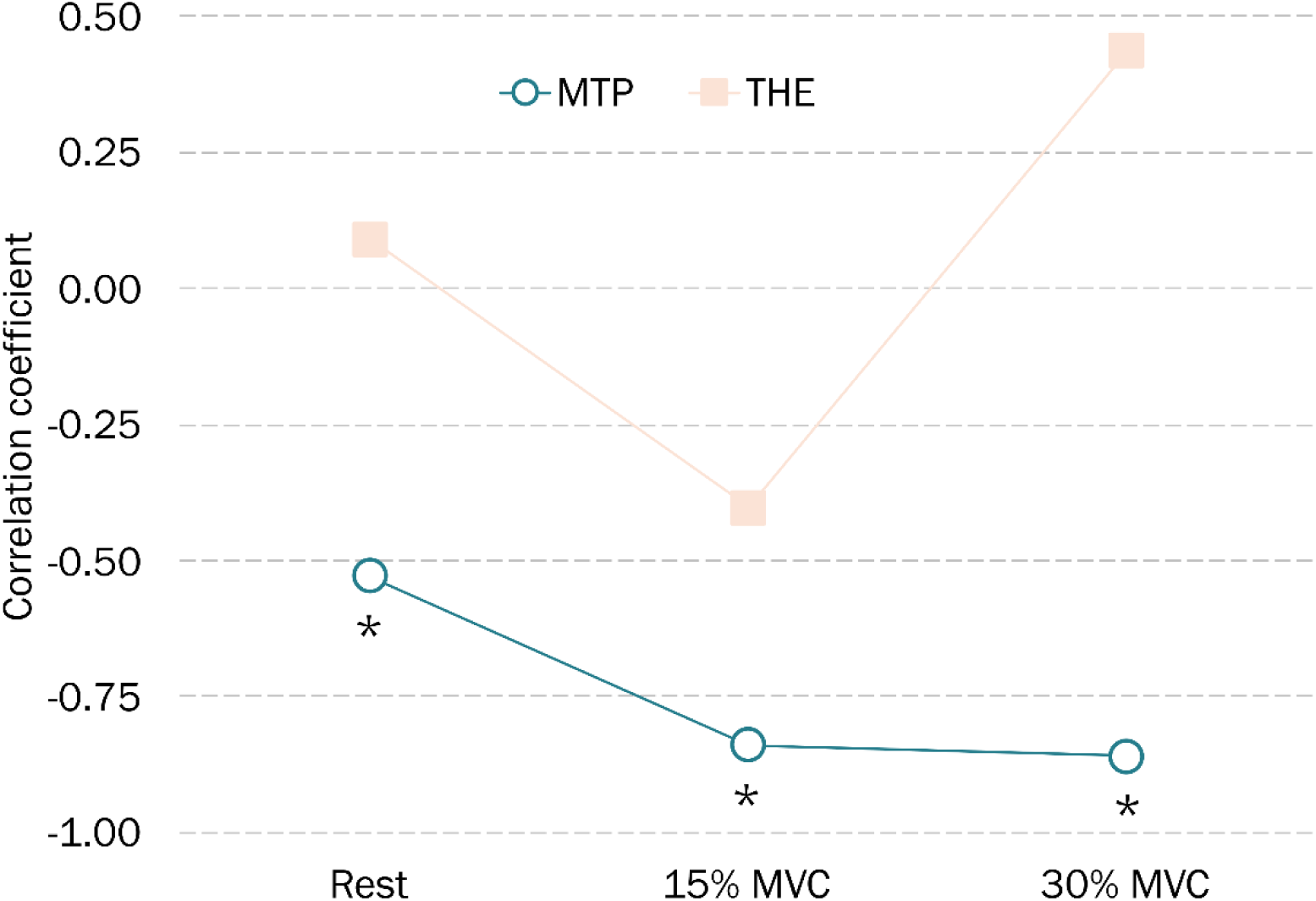
Pearson’s correlation coefficients between skin layer and subcutaneous fat thickness and tissue stiffness estimations of MyotonPRO (MTP) and time-harmonic elastography (THE) stiffness estimations at different contraction intensities. The correlations are significant at the 1% threshold or higher.

### THE Stiffness and Wave Attenuation

Large effects of contraction intensity were found for THE stiffness (Rest: 5.3 [1.2] kPa, 15% MVC: 10.8 [2.2] kPa, 30% MVC: 15.6 [6.1] kPa, *F* = 46.58, p < 0.001, ES = 0.65) and for THE wave attenuation (Rest: 0.83 [0.19] s/m, 15% MVC: 1.26 [0.34] s/m, 30% MVC: 1.42 [0.36] s/m, *F* = 30.13, p < 0.001, ES = 0.55). The correlation values between the different outcomes of the two technologies at the resting muscle state, 15% MVC, 30% MVC are presented in Tables 1-3.

**Table 1.**
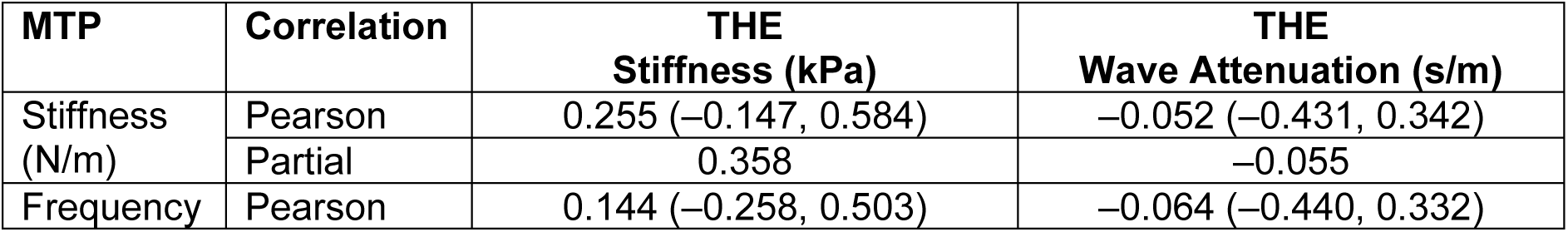
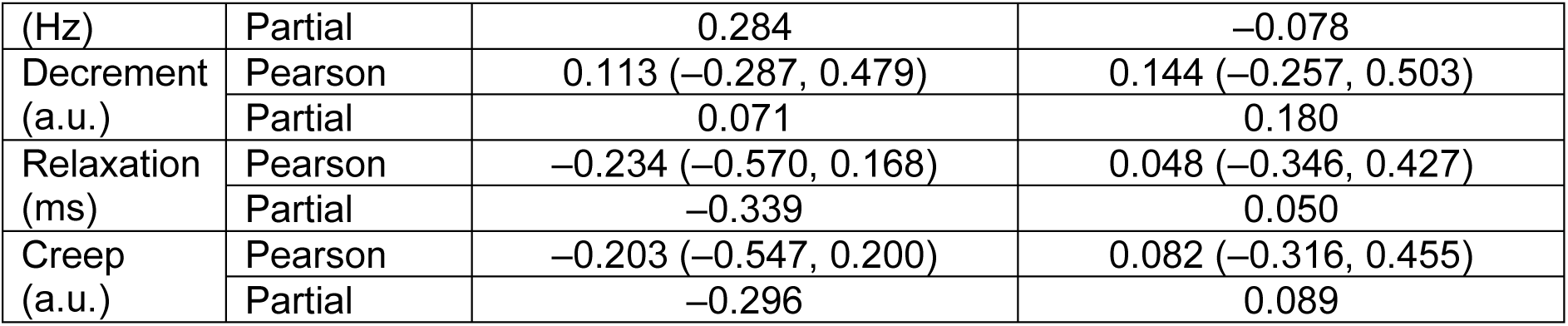
Strength of the relationships as Pearson’s (with lower, upper 95% confidence bounds) and partial (with adjustment of SLSF thickness) correlation coefficients at resting muscle state (**0% MVC**).

**Table 2.** Strength of the relationships as Pearson’s (with lower, upper 95% confidence bounds) and partial (with adjustment of SLSF thickness) correlation coefficients at **15% MVC**.

| MTP | Correlation | THE Stiffness (kPa) | THE Wave Attenuation (s/m) |
| --- | --- | --- | --- |
| Stiffness (N/m) | Pearson | 0.294 (-0.105, 0.612) | -0.316 (-0.627, 0.081) |
|  | Partial | -0.087 | -0.181 |
| Frequency (Hz) | Pearson | 0.273 (-0.128, 0.597) | -0.335 (-0.639, 0.060) |
|  | Partial | -0.159 | -0.220 |
| Decrement (a.u.) | Pearson | -0.064 (-0.441, 0.331) | 0.367 (-0.024, 0.660) |
|  | Partial | **0.511** | 0.274 |
| Relaxation (ms) | Pearson | -0.351 (-0.650, 0.042) | 0.284 (-0.116, 0.605) |
|  | Partial | -0.042 | 0.124 |
| Creep (a.u.) | Pearson | -0.354 (-0.652, 0.039) | 0.287 (-0.113, 0.607) |
|  | Partial | -0.047 | 0.128 |

**Table 3.** Strength of the relationships as Pearson’s (with lower, upper 95% confidence bounds) and partial (with adjustment of SLSF thickness) correlation coefficients at **30% MVC**.

| MTP | Correlation | THE Stiffness (kPa) | THE Wave Attenuation (s/m) |
| --- | --- | --- | --- |
| Stiffness (N/m) | Pearson | -0.547 (-0.771, -0.202) | 0.306 (-0.092, 0.620) |
|  | Partial | -0.373 | 0.232 |
| Frequency (Hz) | Pearson | -0.537 (-0.765, -0.189) | 0.226 (-0.177, 0.564) |
|  | Partial | -0.352 | 0.070 |
| Decrement (a.u.) | Pearson | 0.369 (-0.021, 0.662) | 0.013 (-0.377, 0.398) |
|  | Partial | 0.125 | 0.212 |
| Relaxation (ms) | Pearson | 0.484 (0.119, 0.734) | -0.344 (-0.646, 0.050) |
|  | Partial | **0.236** | -0.309 |
| Creep (a.u.) | Pearson | 0.488 (0.124, 0.736) | -0.333 (-0.638, 0.062) |
|  | Partial | **0.245** | -0.284 |

### MTP Outcomes

Large effects of contraction intensity were found for MTP stiffness (Rest: 337.3 [49.3] N/m, 15% MVC: 449.5 [115.8] N/m, 30% MVC: 529.4 [160.7] N/m, *F* = 49.25, p < 0.001, ES = 0.66) and for the MTP oscillation frequency (Rest: 17.2 [2.1] Hz, 15% MVC: 18.9 [3.4] Hz, 30% MVC: 19.5 [4.1] Hz, *F* = 13.94, p < 0.001, ES = 0.39).

MTP logarithmic decrement showed a moderate effect of contraction intensity (Rest: 0.90 [0.10], 15% MVC: 0.85 [0.13], 30% MVC: 0.88 [0.16], *F* = 3.74, p < 0.05, ES = 0.13).

Large effects of contraction intensity were also found for MTP relaxation time (Rest: 16.7 [2.7] ms, 15% MVC: 11.9 [3.2] ms, 30% MVC: 10.6 [3.3] ms, *F* = 147.75, p < 0.001, ES = 0.86) and for the MTP Creep (Rest: 1.04 [0.16], 15% MVC: 0.74 [0.17], 30% MVC: 0.66 [0.18], *F* = 178.67, p < 0.001, ES = 0.88) with decreasing values while increasing intensity.

Interestingly, at 30% MVC the THE stiffness values were inversely associated with MTP stiffness results (r = –0.547, p < 0.01, Figure 4). The strength of the relationship between both quantities changed (r = –0.373, p = 0.07) after controlling for participants’ SLSF thickness.

**Figure 4.**
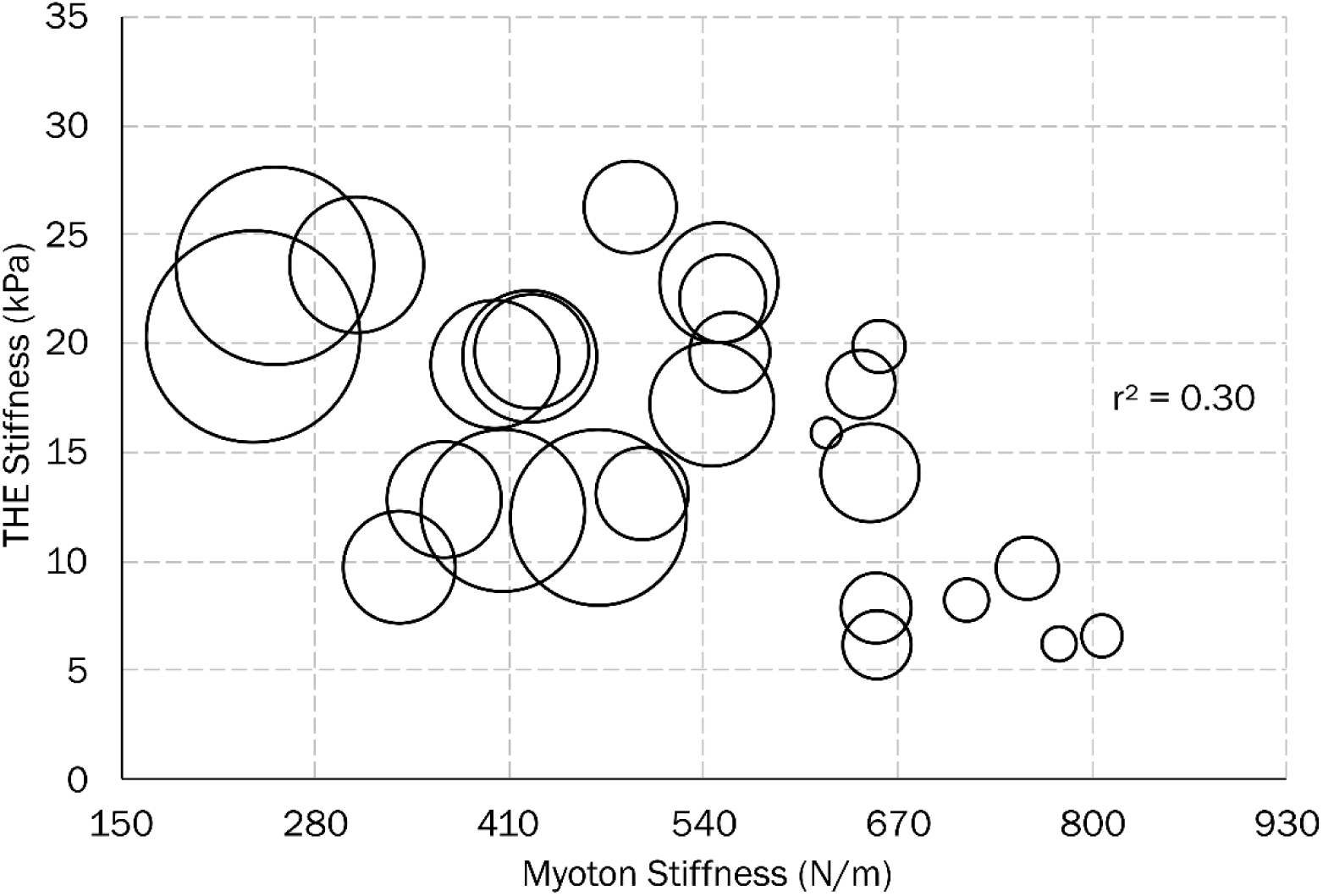
THE stiffness over MTP stiffness where the size of the circles indicates skin layer and subcutaneous fat thickness.

**Figure 5.**
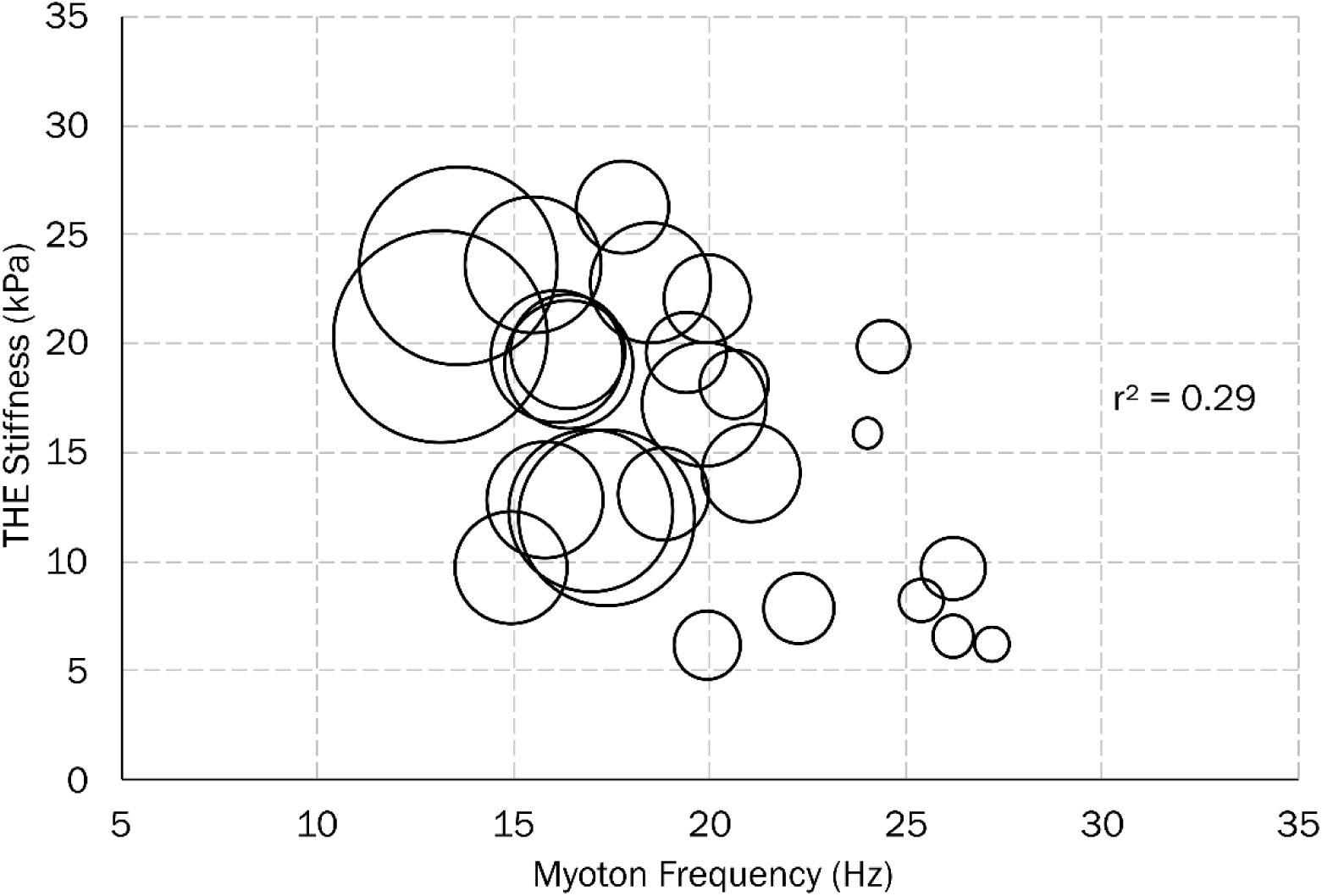
THE stiffness over MTP oscillation frequency where the size of the circles indicates skin layer and subcutaneous fat thickness.

**Figure 6.**
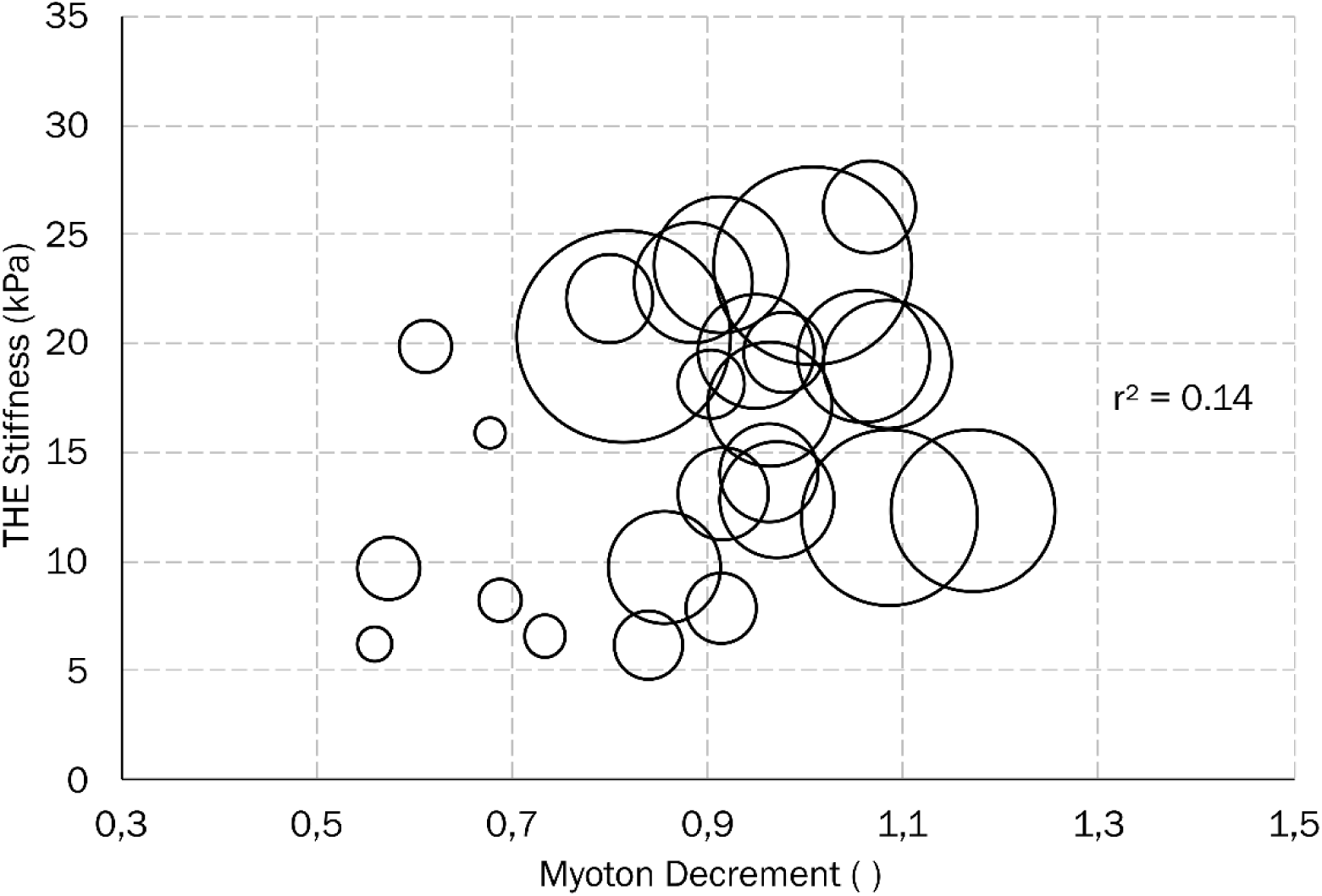
THE stiffness over MyotonPRO logarithmic decrement where the size of the circles indicates skin layer and subcutaneous fat thickness.

**Figure 7.**
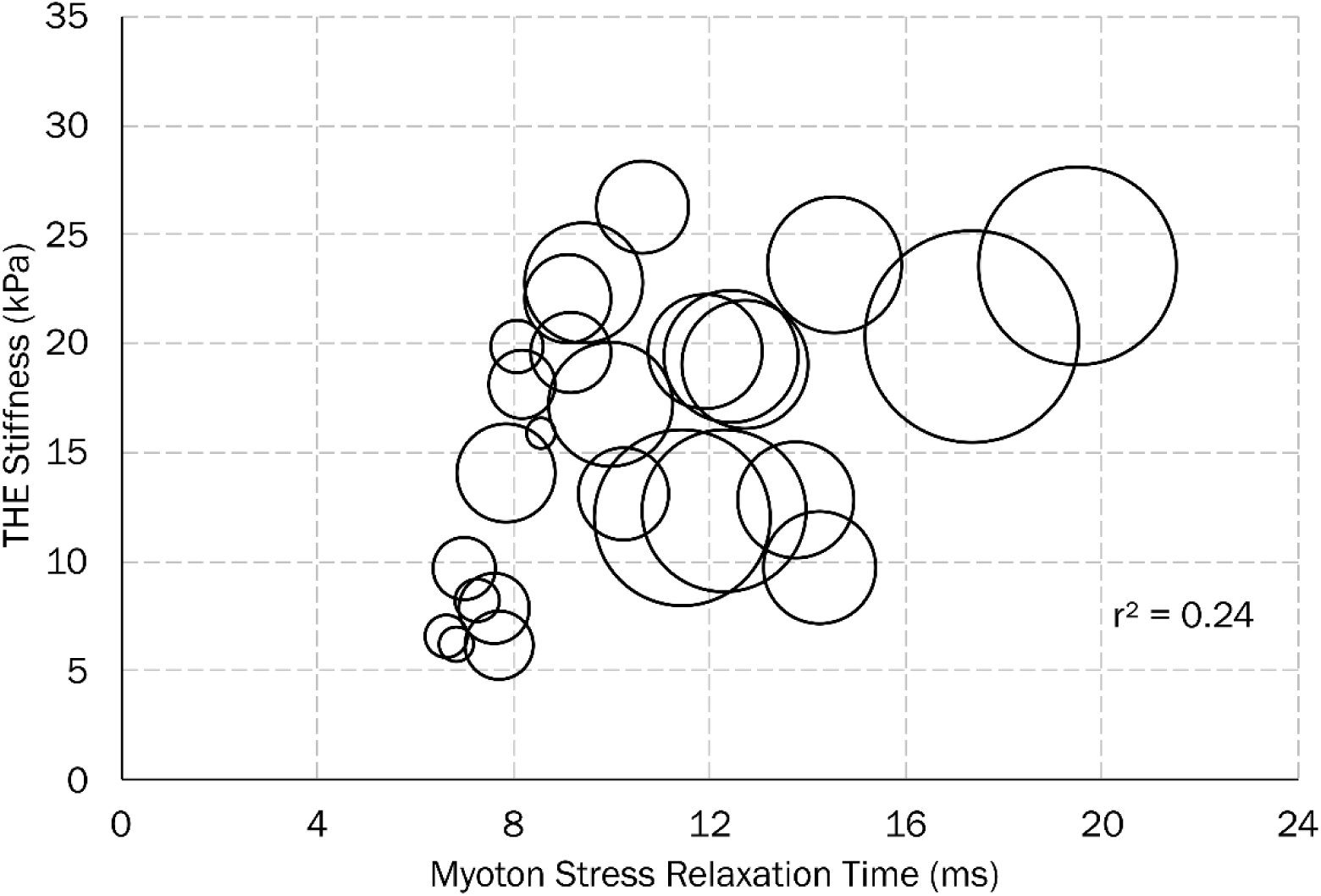
THE stiffness over MyotonPRO relaxation time where the size of the circles indicates skin layer and subcutaneous fat thickness.

**Figure 8.**
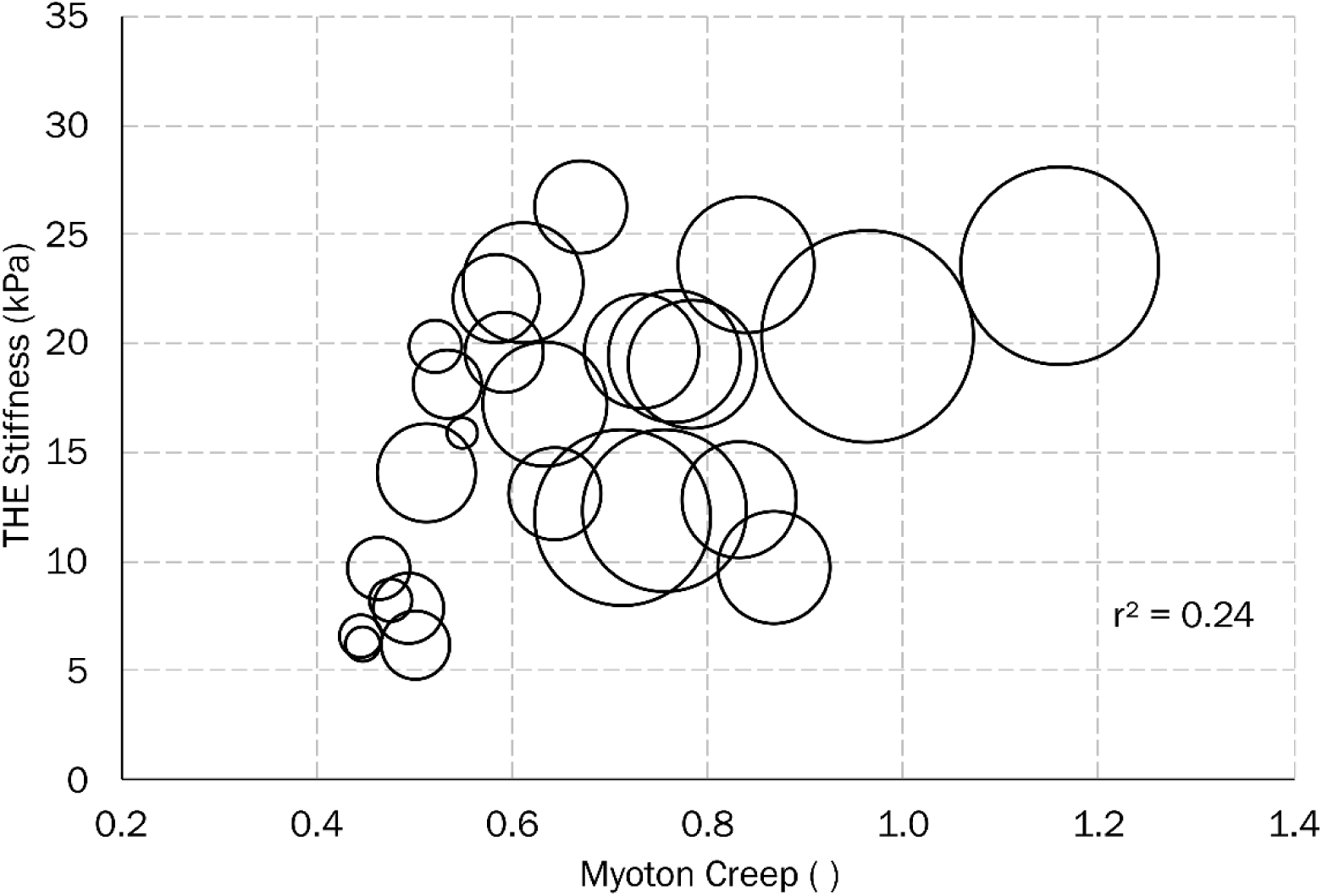
THE stiffness over MyotonPRO creep where the size of the circles indicates skin layer and subcutaneous fat thickness.

## Discussion

### Principal Findings

Both THE and MTP detected contraction intensity-dependent changes across all measured outcomes, confirming that each technology is sufficiently sensitive to distinguish resting muscle from actively contracting muscle at 15% and 30% MVC. This is consistent with prior work showing that shear wave speed in the vastus lateralis increases roughly linearly with force output [17, 18], and that MTP stiffness and oscillation frequency respond to voluntary activation in a dose-dependent fashion [1].

Despite this shared sensitivity, the two modalities did not track each other closely. Correlations between MTP and THE outcomes were weak to moderate at rest and became inconsistent or inverted at higher contraction levels. At 30% MVC, MTP stiffness was significantly and inversely correlated with THE stiffness (r = −0.55), and this relationship was only partly explained by SLSF thickness (partial r = −0.37). These findings indicate that MTP and THE are not measuring the same aspect of muscle mechanics, even when applied to the same muscle under the same contraction conditions. From a practical standpoint, results from the two technologies cannot be assumed to be interchangeable, particularly during active muscle contraction.

### Subcutaneous Tissues

Subcutaneous fat thickness was significantly and negatively correlated with MTP stiffness and oscillation frequency at all contraction levels (Figure 3), whereas its effect on THE stiffness was comparatively weaker. This divergence reflects a fundamental difference in measurement physics: MTP’s 0.40 N mechanical impulse must propagate through all overlying tissue layers (skin, adipose, and fascia) before reaching the target muscle, and each layer absorbs and attenuates the oscillation signal in proportion to its thickness and compliance. As a result, individuals with greater SLSF thickness systematically show lower MTP stiffness readings, independently of the actual state of the underlying muscle. This “fat buffering” effect has been documented in previous studies, with reported correlations between SLSF thickness and MTP stiffness ranging from r = −0.28 to r = −0.84 across different muscles [19-21]. A recent study comparing MTP and shear wave elastography in the rectus femoris reached essentially the same conclusion as the present work: MTP and SWE are not interchangeable in muscle stiffness assessment, primarily because subcutaneous adipose tissue confounds the MTP signal in ways that do not affect depth-resolved elastography [22].

THE stiffness, by contrast, is derived from shear wave propagation speed, an intensive material property that is not accumulated along the tissue path above the region of interest. In the present study, the THE ROI was defined to span 0 to 20 mm depth, co-located with the MTP probe, and therefore encompassed the same superficial tissue layers. Despite this, THE stiffness showed weaker and non-significant correlations with SLSF thickness compared to MTP (Figure 3). This divergence reflects a difference in measurement physics: the MTP oscillatory response is attenuated by soft, compliant superficial layers through which the mechanical impulse must propagate, whereas THE estimates local shear wave speed at each position within the wave field, which is not governed by the same cumulative attenuation mechanism. These findings suggest that SLSF thickness represents a fundamental source of disparity between the two modalities that increases with contraction intensity as the mechanical contrast between muscle and overlying adipose tissue increases. As the muscle contracts and stiffens, the absolute mechanical contrast between the muscle and the overlying adipose layer increases, and the proportion of the MTP signal attributable to non-muscle tissue increases relative to the contribution of the muscle.

### Probe Orientation of the MTP Device

The MTP device incorporates a tri-axial accelerometer that enforces a perpendicularity tolerance of ±5° and rejects measurements that exceed this window [23, 24]. In the present study, measurements were applied over the distal portion of the vastus lateralis with the probe held perpendicular to the skin, consistent with the guidelines of the manufacturer. However, the vastus lateralis has an oblique fiber pennation angle (typically 12–20°), which means that even a perfectly perpendicular probe placement creates a non-trivial angle between the probe axis and the dominant muscle fiber direction. This geometric mismatch has implications for both MTP and THE. For MTP, the measured MTP stiffness reflects a composite response that blends the anisotropic mechanical properties of tissue weighted by fiber orientation and depth distribution. For THE, the propagation direction of the induced shear waves relative to fiber orientation determines whether the measurement is sampling the fiber-parallel or fiber-perpendicular shear modulus, values that differ by a factor of 2–3 in active skeletal muscle [11, 25]. The directional filtering applied in the k-MDEV pipeline (centered at 0° and 180°) selectively emphasizes horizontally propagating waves, which in our measurement geometry were oriented predominantly perpendicular to the longitudinal axis of muscle and partly oblique to the fiber direction. Both modalities are therefore sampling a direction-dependent projection of the mechanical anisotropy of tissue, but they do so differently, and this may contribute to the weak and inconsistent cross-modal correlations observed. Future studies should systematically vary probe orientation relative to fiber architecture (ideally guided by real-time B-mode imaging of fiber angle) to disentangle anisotropy effects from other sources of modality divergence.

### Frequency-Dependent Viscoelasticity

A third, and perhaps underappreciated, source of divergence is the operating frequency of each device. Skeletal muscle is a viscoelastic material whose apparent stiffness increases with the frequency of mechanical excitation, a phenomenon known as viscoelastic dispersion. The MTP characterizes tissue at its natural resonant frequency, which in the vastus lateralis under the conditions of the present study ranged from approximately 17 to 20 Hz. THE used superimposed frequencies of 60, 70, and 80 Hz. These two ranges sample different positions on the frequency-stiffness spectrum of tissue, and the difference is not trivial: rheometric studies of soft biological tissues have reported stiffness increases of 50–120% over frequency ranges spanning 10 to 100 Hz [26, 27]. This means that THE will systematically report higher absolute stiffness values than MTP for the same tissue state because they are measuring at different operating points of a nonlinear, frequency-dependent mechanical system. Furthermore, during active muscle contraction the viscous (loss) component of the mechanical response increases disproportionately [9, 28], which means the viscosity-induced upshift is larger at contraction than at rest. This provides a mechanistic explanation for why MTP relaxation time and creep showed their strongest associations with THE stiffness at 30% MVC: as contraction deepens the viscous state of the tissue, the inherent sensitivity of MTP to viscoelastic damping begins to carry information about the same underlying mechanical changes that THE resolves through shear wave speed dispersion.

### The Inverse Relationship at 30% MVC

The significant inverse correlation between MTP Stiffness and THE Stiffness at 30% MVC (r = −0.55, p < 0.01; Figure 4) deserves careful interpretation, because it appears counterintuitive: a device reporting higher stiffness values is associated with a device reporting lower values in the same muscle at the same time. Several converging mechanisms can explain this without requiring a fundamental measurement error in either device. First, participants with thicker SLSF layers (who exhibit artificially low MTP stiffness due to fat buffering) may also have larger absolute muscle volumes that produce higher shear wave speeds when contracting at the same relative force level (30% of their personal MVC). This creates a negative cross-participant correlation through body composition. Second, at high contraction levels the MTP’s 0.40 N impulse becomes small relative to the increased stiffness of tissue, potentially causing partial saturation or floor effects in the oscillation amplitude, while THE continues to track shear wave speed increases linearly. Third, controlling for SLSF thickness partially attenuated the inverse relationship (partial r = −0.37, p = 0.07), consistent with body composition being a major mediating variable. These explanations are not mutually exclusive, and it is likely that all three contribute in proportion to individual variation in body composition and muscle architecture. The practical implication is clear: cross-sectional comparisons of muscle stiffness between individuals should not mix MTP and THE values without explicitly accounting for SLSF thickness, and within-protocol consistency should be prioritized.

### Comparison between MTP Stiffness and THE Stiffness

It would be a mistake, however, to conclude from the weak cross-modal correlations that one technology is superior to the other. They measure different things, and both things are scientifically meaningful. THE stiffness provides a spatially resolved, geometry-independent shear wave speed that reflects the intrinsic elastic state of the muscle fibers and their connective tissue matrix at the specific depth and location selected. This makes it well-suited for quantifying regional heterogeneity, comparing muscles with different geometries, and detecting pathological stiffening in specific tissue layers. MTP stiffness, relaxation time, and creep capture a composite mechanical response that integrates viscoelastic behavior across tissue layers at the timescale of the impulse response. This may actually be an advantage for some applications: the integrated nature of the signal could be sensitive to systemic changes in tissue tone (such as those induced by fatigue, delayed-onset muscle soreness, or spasticity) that manifest diffusely rather than in a single layer. MTP relaxation time and creep showed their best associations with THE stiffness at 30% MVC, not at rest, which is consistent with the idea that during active contraction the viscoelastic state of the muscle becomes the dominant shared signal between the two modalities. Future work might productively use both methods together (THE to anchor absolute stiffness values and MTP to capture rapid longitudinal changes in a field setting) rather than treating them as competing alternatives.

### Limitations

Several limitations should be considered when interpreting these results. The study included only healthy young adults with a relatively narrow range of body mass indices, which limits generalizability to clinical populations where both SLSF thickness and muscle pathology may vary more widely. The low contraction levels tested (15% and 30% MVC) were chosen to ensure stable, controllable force production during imaging, but the behavior at higher contraction intensities (where MTP signal saturation effects might become more pronounced) remains unexplored. The THE processing used a fixed tissue density assumption of 1000 kg/m³ and isotropic reconstruction; anisotropy-aware inversion could yield more accurate stiffness maps in the pennate fiber architecture of the vastus lateralis [29]. Finally, the cross-sectional design prevents causal inference: we can describe the relationship between the two modalities, but not identify whether a given discrepancy reflects a measurement artifact or a genuine difference in the mechanical quantity each device is sensitive to. Longitudinal intervention studies (comparing both devices before and after training, immobilization, or pathology) would help clarify this.

In summary, MyotonPRO and time-harmonic elastography are both sensitive to contraction-induced mechanical changes in the vastus lateralis, but they measure fundamentally different quantities and their outputs cannot be used interchangeably. MTP Stiffness is an extensive, geometry-dependent quantity that integrates mechanical contributions from all tissue layers within the probe’s depth of influence, with the response weighted exponentially toward more superficial structures. The shear modulus derived from THE is an intensive material property estimated from propagating wave speed, which is geometry-independent by definition regardless of the spatial extent of the analysis ROI. Although the THE ROI in this study was constrained to match the approximate depth and lateral extent of the MTP measurement footprint (0–20 mm depth, 6 mm width) to maximize comparability, this design choice does not eliminate the fundamental differences between the two modalities: the physical quantities they measure are distinct (N/m versus kPa), the frequencies at which tissue is interrogated differ substantially (approximately 17–20 Hz for MTP versus 60–80 Hz for THE), and the depth-weighting structures within the shared volume are governed by different physical mechanisms. These differences have direct consequences for how results from each device should be interpreted, particularly in the presence of individual variation in subcutaneous tissue composition.

At rest, the two modalities share a weak positive statistical relationship driven primarily by tissue stiffness. During active contraction, the relationship degrades and can invert, with subcutaneous fat thickness acting as a key confounding variable that suppresses MTP readings while leaving THE measurements relatively unaffected. Researchers and clinicians who use these devices should explicitly account for SLSF thickness when interpreting results and should not assume that a change detected by one technology implies a corresponding change detectable by the other.

Rather than viewing MTP and THE as competing alternatives, they are better understood as complementary tools: MTP is fast, portable, and sensitive to composite viscoelastic state across tissue layers, making it well-suited for longitudinal monitoring and field-based screening; THE provides quantitative, tissue-layer-specific stiffness maps that are appropriate for diagnostic imaging, research validation, and applications requiring isolation of intrinsic muscle mechanical properties. Used together with an awareness of their differing measurement physics, the two modalities can offer a richer characterization of neuromuscular mechanics than either alone.

## Acknowledgements

We are sincerely grateful to Mr. Moritz Waage and Mr. Matti Panian for their invaluable support in participant recruitment and for their assistance throughout the data collection process. We would also like to acknowledge financial support from the Federal Ministry for Economic Affairs and Energy (KK5000503BM4 - MUSKEL). The sponsor had no role in the study design, data collection, analysis, or interpretation.

## Statements and Declarations

### Author Contributions

**EK**: Conceptualization, Data curation, Writing - original draft, Writing - review & editing, Visualization, Investigation, Formal analysis, Methodology, Project administration and Software. **GV**: Writing - review & editing, Formal analysis, Methodology. **TM**: Writing - review & editing, Formal analysis, Methodology. **SP**: Conceptualization, Data curation, Writing - review & editing, Investigation. **RS**: Conceptualization, Funding acquisition, Writing - review & editing, Supervision. **TB**: Conceptualization, Funding acquisition, Writing - review & editing. **KD**: Conceptualization, Funding acquisition, Writing - review & editing, Supervision. **IS**: Conceptualization, Funding acquisition, Writing - review & editing, Supervision. **HA**: Conceptualization, Data curation, Writing - original draft, Writing - review & editing, Visualization, Investigation, Formal analysis, Methodology, Project administration and Software, Supervision.

### Data Availability

The datasets generated and/or analyzed during the current study are not publicly available due to ethical restrictions and participant privacy.

### Code Availability

Custom code used for data processing and analysis is not publicly available but can be shared with researchers upon reasonable request.

## Appendix A

## A1. MyotonPRO: Damped Mass-Spring Oscillator Model

The probe-tissue system is modeled as a damped harmonic oscillator with an effective mass of ≈ 18 g. Following a brief mechanical impulse (0.40 N, 15 ms), the probe oscillates at the natural resonant frequency of tissue, and MTP stiffness (*S*) and oscillation frequency *F*) are extracted from the free decay acceleration signal:

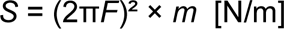

MTP stiffness is an extensive (geometry-dependent) quantity. It integrates mechanical contributions from all tissue layers within approximately 15–20 mm of depth, including skin, subcutaneous fat, fascia, and muscle. Because it depends on both material properties and the effective contact geometry, it cannot be directly equated to an intrinsic material modulus [6].

## A2. Time-Harmonic Elastography: Shear Wave Propagation

Time-harmonic elastography (THE) applies continuous harmonic vibration (60, 70, 80 Hz) to generate shear waves whose propagation speed is determined by the intrinsic shear modulus of tissue. THE stiffness (*G*) is recovered via k-MDEV inversion using local phase gradients of the measured wave field:

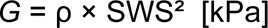

where SWS is the shear wave speed and ρ = 1000 kg/m³. THE stiffness is an intensive (geometry-independent) material property. By selecting a region of interest on the B-mode image, the measurement can be spatially confined to the muscle belly, excluding all superficial tissue layers (Aghamiry et al. 2025, Meyer et al. 2025). THE wave attenuation is simultaneously derived from the imaginary component of the complex wavenumber, providing a complementary measure of viscoelastic energy dissipation.

## A3. Why Direct Conversion Between MTP Stiffness and THE Stiffness Is Not Feasible

Four structural reasons prevent reliable conversion between MTP Stiffness and THE Stiffness.

### Geometry dependence

MTP Stiffness depends on both material modulus and effective contact geometry: *S* = *G* × *A* / *L* × *g*, where *A* is the effective contact area, *L* is the characteristic depth, and *g* is a geometric correction factor [6]. None of these are known *in vivo*. THE stiffness directly recovers *G* without geometric assumptions.

Depth **integration vs. spatial localization**: MTP stiffness represents an exponentially depth-weighted composite of all layers from surface to ∼20 mm. Subcutaneous adipose tissue thickness is negatively correlated with MTP stiffness (*r* = –0.47 to – 0.84; Fröhlich-Zwahlen et al. 2014, Kelly et al. 2018), meaning fat content confounds deep-muscle assessment. THE stiffness can be spatially confined to a user-defined ROI within the muscle belly, excluding all overlying layers.

Frequency **dispersion**: MTP operates at the natural resonant frequency of tissue (∼10–50 Hz), while THE uses controlled frequencies of 60–80 Hz. Because soft tissue stiffness increases with excitation frequency (10–120% over this range; Chen et al. 2009, Nicolle et al. 2023), the two modalities inherently sample different regions of the viscoelastic spectrum. Bridging this gap requires explicit viscoelastic modeling with tissue-specific parameters that are not available in standard clinical measurements [30].

Mechanical **anisotropy**: Skeletal muscle shear modulus measured parallel to fibers exceeds that perpendicular to fibers by a factor of 2–3 [29, 31]. MTP probes predominantly perpendicular to the skin surface at an anatomically variable angle to fiber direction. THE samples shear wave propagation along a fixed imaging plane. Both instruments therefore measure direction-weighted projection of an anisotropic mechanical tensor, but the projection angles differ and are not controlled independently. Empirical attempts to relate the two modalities have yielded only modest agreement (*R*² = 0.21–0.30; Feng et al. 2018), with substantial unexplained variance even under controlled laboratory conditions.

## References

1. Aird L, Samuel D, Stokes M. Quadriceps muscle tone, elasticity and stiffness in older males: reliability and symmetry using the MyotonPRO. Arch Gerontol Geriatr. 2012;55(2):e31–9. doi: 10.1016/j.archger.2012.03.005.

2. Gavronski G, Veraksits A, Vasar E, Maaroos J. Evaluation of viscoelastic parameters of the skeletal muscles in junior triathletes. Physiol Meas. 2007;28(6):625–37. doi: 10.1088/0967-3334/28/6/002.

3. Tzschätzsch H, Nguyen Trong M, Scheuermann T, Ipek-Ugay S, Fischer T, Schultz M, et al. Two-dimensional time-harmonic elastography of the human liver and spleen. Ultrasound Med Biol. 2016;42(11):2562–71. doi: 10.1016/j.ultrasmedbio.2016.07.004.

4. Aghamiry HS, Meyer T, Klemmer Chandia S, Engl P, Valli G, Wu Y, et al. Muscle activation assessment using ultrasound time-harmonic elastography and tonic vibration reflex. J Biomech. 2026.

5. Feng YN, Li YP, Liu CL, Zhang ZJ. Assessing the elastic properties of skeletal muscle and tendon using shearwave ultrasound elastography and MyotonPRO. Sci Rep. 2018;8(1):17064. doi: 10.1038/s41598-018-34719-7.

6. Zhang M, Zheng YP, Mak AF. Estimating the effective Young’s modulus of soft tissues from indentation tests - nonlinear finite element analysis of effects of friction and large deformation. Med Eng Phys. 1997;19(6):512–7. doi: 10.1016/s1350-4533(97)00017-9.

7. Snedeker JG, Niederer P, Schmidlin FR, Farshad M, Demetropoulos CK, Lee JB, et al. Strain-rate dependent material properties of the porcine and human kidney capsule. J Biomech. 2005;38(5):1011–21. doi: 10.1016/j.jbiomech.2004.05.036.

8. Chen S, Urban MW, Pislaru C, Kinnick R, Zheng Y, Yao A, et al. Shearwave dispersion ultrasound vibrometry (SDUV) for measuring tissue elasticity and viscosity. IEEE Trans Ultrason Ferroelectr Freq Control. 2009;56(1):55–62. doi: 10.1109/TUFFC.2009.1005.

9. Klatt D, Papazoglou S, Braun J, Sack I. Viscoelasticity-based MR elastography of skeletal muscle. Phys Med Biol. 2010;55(21):6445–59. doi: 10.1088/0031-9155/55/21/007.

10. Mullix J, Warner M, Stokes M. Testing muscle tone and mechanical properties of rectus femoris and biceps femoris using a novel hand held MyotonPRO device: relative ratios and reliability. Work Pap Health Sci. 2012;1(1):1–8.

11. Meyer T, Wellge B, Barzen G, Klemmer Chandia S, Knebel F, Hahn K, et al. Cardiac elastography with external vibration for quantification of diastolic myocardial stiffness. J Am Soc Echocardiogr. 2025;38(5):431–42. doi: 10.1016/j.echo.2024.11.009.

12. Felsberg M, Sommer G. The monogenic signal. IEEE Trans Signal Process. 2001;49(12):3136–44. doi: 10.1109/78.969520.

13. Unser M, Sage D, Van De Ville D. Multiresolution monogenic signal analysis using the Riesz-Laplace wavelet transform. IEEE Trans Image Process. 2009;18(11):2402–18. doi: 10.1109/TIP.2009.2027628.

14. Schneider S, Peipsi A, Stokes M, Knicker A, Abeln V. Feasibility of monitoring muscle health in microgravity environments using Myoton technology. Med Biol Eng Comput. 2015;53(1):57–66. doi: 10.1007/s11517-014-1211-5.

15. Agyapong-Badu S, Warner M, Samuel D, Stokes M. Measurement of ageing effects on muscle tone and mechanical properties of rectus femoris and biceps brachii in healthy males and females using a novel hand-held myometric device. Arch Gerontol Geriatr. 2016;62:59–67. doi: 10.1016/j.archger.2015.09.011.

16. Ditroilo M, Hunter AM, Haslam S, De Vito G. The effectiveness of two novel techniques in establishing the mechanical and contractile responses of biceps femoris. Physiol Meas. 2011;32(8):1315–26. doi: 10.1088/0967-3334/32/8/020.

17. Shinohara M, Sabra K, Gennisson JL, Fink M, Tanter M. Real-time visualization of muscle stiffness distribution with ultrasound shear wave imaging during muscle contraction. Muscle Nerve. 2010;42(3):438–41. doi: 10.1002/mus.21723.

18. Yang Y, Shahryari M, Meyer T, Marticorena Garcia SR, Görner S, Salimi Majd M, et al. Explorative study using ultrasound time-harmonic elastography for stiffness-based quantification of skeletal muscle function. Z Med Phys. 2025;35(4):470–82. doi: 10.1016/j.zemedi.2024.03.001.

19. Kelly JP, Koppenhaver SL, Michener LA, Proulx L, Bisagni F, Cleland JA. Characterization of tissue stiffness of the infraspinatus, erector spinae, and gastrocnemius muscle using ultrasound shear wave elastography and superficial mechanical deformation. J Electromyogr Kinesiol. 2018;38:73–80. doi: 10.1016/j.jelekin.2017.11.001.

20. Fröhlich-Zwahlen AK, Casartelli NC, Item-Glatthorn JF, Maffiuletti NA. Validity of resting myotonometric assessment of lower extremity muscles in chronic stroke patients with limited hypertonia: a preliminary study. J Electromyogr Kinesiol. 2014;24(5):762–9. doi: 10.1016/j.jelekin.2014.06.007.

21. Bravo-Sanchez A, Abian P, Sanchez-Infante J, Esteban-Gacia P, Jimenez F, Abian-Vicen J. Objective assessment of regional stiffness in vastus lateralis with different measurement methods: a reliability study. Sensors (Basel). 2021;21(9):3213. doi: 10.3390/s21093213.

22. Ley CD, Martinez Valdes E, Murtagh CF, Power J, Nobes L, Drust B. MyotonPRO is not comparable to shear wave elastography in the measurement of rectus femoris muscle stiffness due to interference of subcutaneous adipose tissue. Scand J Med Sci Sports. 2025;35(8):e70095. doi: 10.1111/sms.70095.

23. Astrand AP, Jalkanen V, Andersson BM, Lindahl OA. Contact angle and indentation velocity dependency for a resonance sensor - evaluation on soft tissue silicone models. J Med Eng Technol. 2013;37(3):185–96. doi: 10.3109/03091902.2013.773097.

24. Muckelt PE, Warner MB, Cheliotis-James T, Muckelt R, Hastermann M, Schoenrock B, et al. Protocol and reference values for minimal detectable change of MyotonPRO and ultrasound imaging measurements of muscle and subcutaneous tissue. Sci Rep. 2022;12(1):13654. doi: 10.1038/s41598-022-17507-2.

25. Gennisson JL, Deffieux T, Mace E, Montaldo G, Fink M, Tanter M. Viscoelastic and anisotropic mechanical properties of in vivo muscle tissue assessed by supersonic shear imaging. Ultrasound Med Biol. 2010;36(5):789–801. doi: 10.1016/j.ultrasmedbio.2010.02.013.

26. Kiss MZ, Varghese T, Hall TJ. Viscoelastic characterization of in vitro canine tissue. Phys Med Biol. 2004;49(18):4207–18. doi: 10.1088/0031-9155/49/18/002.

27. Nicolle S, Palierne JF, Mitton D, Follet H, Confavreux CB. Multi-frequency shear modulus measurements discriminate tumorous from healthy tissues. J Mech Behav Biomed Mater. 2023;140:105721. doi: 10.1016/j.jmbbm.2023.105721.

28. Schrank F, Warmuth C, Görner S, Meyer T, Tzschätzsch H, Guo J, et al. Real-time MR elastography for viscoelasticity quantification in skeletal muscle during dynamic exercises. Magn Reson Med. 2020;84(1):103–14. doi: 10.1002/mrm.28095.

29. Meyer T, Klemmer Chandia S, Engl P, Valli G, Wu Y, Jenderka K, et al. Shear-wave anisotropy of the vastus lateralis during low-level isometric contraction measured with ultrasound time-harmonic elastography. medRxiv. 2025. doi: 10.1101/2025.09.30.25336813.

30. Lakes R. Viscoelastic materials. Cambridge: Cambridge University Press; 2009.

31. Eby SF, Song P, Chen S, Chen Q, Greenleaf JF, An KN. Validation of shear wave elastography in skeletal muscle. J Biomech. 2013;46(14):2381–7. doi: 10.1016/j.jbiomech.2013.07.033.

